# Beyond ratios - flexible and resilient nurse staffing options to deliver cost-effective hospital care and address staff shortages: a simulation and economic modelling study

**DOI:** 10.1101/2020.11.30.20240945

**Authors:** Peter Griffiths, Christina Saville, Jane E Ball, Jeremy Jones, Thomas Monks, On behalf of the Safer Nursing Care Tool study team

## Abstract

**Background:** In the face of pressure to contain costs and make best use of scarce nurses, flexible staff deployment (floating staff between units and temporary hires) guided by a patient classification system may appear an efficient approach to meeting variable demand for care in hospitals.

**Objectives:** We modelled the cost-effectiveness of different approaches to planning baseline numbers of nurses to roster on general medical/surgical units while using flexible staff to respond to fluctuating demand.

**Design and Setting:** We developed an agent-based simulation model, where hospital inpatient units move between being understaffed, adequately staffed or overstaffed as staff supply and demand, measured by a classification system (the Safer Nursing Care Tool) varies. Staffing shortfalls are addressed first by floating staff from overstaffed units, secondly by hiring temporary staff. We compared a standard staffing plan (baseline rosters set to match average demand) with a higher baseline ‘resilient’ plan set to match higher demand, and a lower baseline ‘flexible’ plan. We varied assumptions about temporary staff availability. We estimated the effect of unresolved low staffing on length of stay and death, calculating cost per life saved.

**Results:** Staffing plans with higher baseline rosters led to higher costs but improved outcomes. Cost savings from low baseline staff largely arose because shifts were left understaffed. With limited temporary staff available, the higher baseline ‘resilient’ staffing plan cost £8,653 per life saved compared to the standard plan. The standard plan cost £13,117 per life saved compared to the low baseline flexible plan.

Cost effectiveness for higher baseline staff was further improved with high temporary staff availability. With unlimited temporary staff, the high-baseline staffing plan cost £3,693 per life saved compared to the standard plan and the standard plan cost £4,520 per life saved compared with the low-baseline plan. Cost-effectiveness of higher baseline staffing was even more favourable when negative effects of high temporary staffing were modelled.

**Conclusion:** Flexible staffing can be guided by shift-by-shift measurement of patient demand, but proper attention must be given to ensure that the baseline number of staff rostered is sufficient. Flexible staffing plans that minimise the number of nurses routinely rostered are likely to harm patients because temporary staff may not be available at short notice. Plans that involve low baseline staff rosters and high use of flexible staff therefore do not represent an efficient or effective use of nurses, whereas higher baseline rosters are more resilient in the face of variation and appear cost-effective.

Study registration: ISRCTN 12307968

**Tweetable abstrac:** Economic simulation model of hospital units shows low baseline staff levels with high use of flexible staff are not cost-effective and don’t solve nursing shortages.

**What is already known?:**

- Because nursing is the largest staff group, accounting for a significant proportion of hospital’s variable costs, unit nurse staffing is frequently the target of cost containment measures
- Staffing decisions need to address both the baseline staff establishment to roster, and how best to respond to fluctuating demand as patient census and care needs vary
- Flexible deployment of staff, including floating staff and using temporary hires, has the potential to reduce expenditure while meeting varying patient need, but high use of temporary staff may be associated with adverse outcomes.

**What this paper adds:**

- Low baseline staff rosters that rely heavily on flexible staff provide cost savings largely because units are often left short staffed, which results in adverse patient outcomes and increased non staff costs.
- A staffing plan set to meet average demand appears to be cost effective compared to a plan with a lower baseline but is still associated with frequent short staffing even when using flexible deployments.
- A staffing plan with a higher baseline, set to meet demand 90% of the time, is more resilient in the face of variation and may be highly cost effective

## Introduction

In the face of pressure to contain costs and to use nursing staff, who are in short supply, as efficiently as possible, it is important to understand how best to plan staffing on hospital units. Key decisions relate to the balance between the baseline staffing level to routinely roster (schedule) and the use of flexible staffing (floats and temporary hires) to meet variation in demand caused by variation in census and the needs of patients. The goal is to ensure that the unit staffing system is able to meet fluctuating demand while avoiding wasteful use of human resources and the associated costs. Flexible approaches to staffing deployment to meet variable demand for care have been advocated as a way of ensuring staffing levels are maintained in the face of nursing shortages (Aiken et al., 2013). Some studies have claimed that flexible staffing plans are superior to fixed plans (Kortbeek et al., 2015) but concerns have been raised about potential adverse effects on quality from high use of temporary staff (e.g.Bae et al., 2015, Bae et al., 2010, Dall’Ora et al., 2019b).

In addressing staff shortages, it is important to test assumptions about efficient and effective staff deployment. In a recent review of staffing tools, we found that there is a dearth of evidence about the performance of staffing methods in practice and, in particular, little evidence of the costs and effectiveness of different approaches to determining nurse staffing requirements (Griffiths et al., 2020a). Although there is some evidence that tools in use can measure demand, it is not clear that they identify an optimal staffing level, nor do tools intrinsically address how to schedule staff in advance to meet anticipated variation in demand.

Managing staffing to address variation in demand is a major challenge. Studies have shown substantial variation in demand for nursing care between different hospital units but also from day to say within a unit (Davis et al., 2014b, Griffiths et al., 2018, Van den Heede et al., 2009). Rather than operating with a high baseline staff to accommodate anticipated peaks, flexible deployment of staff is often assumed to be the most efficient approach to meeting such variable demand but there is a lack of evidence for cost-effectiveness or the appropriate balance between the core establishment and flexible deployments (Dall’ora and Griffiths, 2018). Because the adverse effect of low nurse staffing has been demonstrated in many studies and is now widely accepted, there has been much focus on mandatory staffing policies and minimum staffing ratios (Driscoll et al., 2018, Griffiths et al., 2016, Kane et al., 2007). However, the use of ratios is often considered inflexible and inefficient (Buchan, 2005) and even when such a policy is in operation the challenge remains to ensure the proper balance between permanent staff who are rostered in advance, and flexible staffing in order to maintain the required staffing level as demand varies.

In a previous publication, (Saville et al., 2020a) we explored different staffing policies guided by the Safer Nursing Care Tool (The Shelford Group, 2014). The Safer Nursing Care Tool (often referred to by initials SNCT or as the ‘Shelford Tool’) is a patient classification system. It is used in most English National Health Service Hospitals to guide baseline establishments (that is the number of nurses to employ) and, increasingly, daily staff deployments (Ball et al., 2019) informing decision about redeployment of staff between units (floating) or the hiring of temporary staff from the hospital internal pool (bank) or external staffing agencies. In an extensive literature review we found no evidence to determine the cost effectiveness of different ways to use such tools to guide staffing decisions (Griffiths et al., 2020a).

In our previous study, we used a simulation model to compare a standard staffing plan, following Safer Nursing Care Tool recommendations with baseline staffing set to meet average demand, with two alternatives. Firstly, we considered a staffing plan in which fewer staff are rostered routinely, where the emphasis is on the use of flexible deployments, anticipating that most fluctuations in demand would be met by internal redeployment and use of temporary staff. Secondly, we considered a staffing plan in which the baseline staff to be rostered is set at a level that is higher than the mean and is designed to be sufficient to cope with most peaks in demand, while still using flexible staffing this plan emphasised the resilience of the baseline roster in the face of varying levels of demand.

We found that if the number of staff from the permanent establishment rostered on each shift was set at a low level, costs were reduced, but this apparent efficiency was achieved by leaving many shifts understaffed, largely because of the limited availability of staff to float between units or fulfil short notice requests for additional temporary staff (Saville et al., 2020a). Both the levels of understaffing and cost savings were highly dependent on our assumptions about the availability of temporary staff. When more temporary staff were available, understaffing was less common but consequently cost savings were much reduced. In this paper, we consider this further, extending our models to consider the cost-effectiveness of the different approaches.

## Methods

Using data from an observational study in general inpatient units (wards) of acute care hospitals, we developed a simulation model of demand for unit based inpatient nursing. We used this to test various staffing plans guided by the Safer Nursing Care Tool (Griffiths et al., 2020b, Saville et al., 2020a), simulating the staffing levels achieved on each unit and for each shift in the face of variable demand and variable supply of staff. We then estimated the costs and consequences of the resulting staffing levels in an economic model, including cost per life saved with estimates of the effects of low staffing on length of stay and risk of death derived from a recent study (Griffiths et al., 2018).

### Staffing plans

We considered and compare three staffing plans. In the ‘standard’ plan, a baseline number of staff are rostered (scheduled) to work on each unit, set at a level designed to meet average demand observed on that unit, as measured by a patient classification system. This reflects the typical approach to using staffing tools where the mean average of staffing requirements is used to guide decision-making (Griffiths et al., 2020a).

All staffing plans incorporate flexible staffing guided by the demand presented by the patients on each unit on each day, again measured by a patient classification system. If on any given shift demand for staff on any unit is relatively low (for example because of low census or lower than usual patient acuity), excess staff are available to float other units. If the required hours of nursing care on any given shift exceeds the hours that are available from rostered staff on a unit, then ‘excess’ staff can be floated from another unit within the same broad specialty, defined by the hospitals organisational structure (e.g. general medical / surgical). If it is not possible to make up the shortfall with float staff, then temporary staff can be hired from a pool of internal ‘bank’ staff or an external agency. Bank staff include staff employed by the hospital but without fixed hours or assignments, or staff available to work voluntary overtime and so bank hours includes voluntary overtime.

The first alternative staffing plan aims to make more use of flexible staffing, and so has a lower number of staff on the baseline roster than the standard plan. We term this plan ‘flexible’ because the low baseline staff means that it is anticipated that most variation in demand would require the use of these flexible temporary assignments while baseline rosters are set to meet minimal routinely observed demands.

In the second alternative plan, rosters are set at a higher level than the standard plan, anticipating that most upward variation in demand can be met by the staff who are rostered. Although this plan also uses flexible staffing, we term this plan ‘resilient’ because the emphasis is on having enough staff available to meet variation in needs within the routinely scheduled rosters, without recourse to additional measures. See Table 1 for details.

**Table 1.** Staffing scenarios tested in the simulation model.

| Staffing plans* |  |
| --- | --- |
| low baseline (flexible) | Staff roster set to meet 80% of average demand measured by the SNCT across 20 days, set to provide minimum coverage with high use of flexible staffing. |
| Standard (reference) | Staff roster set to meet average demand measured by the Safer Nursing Care Tool (SNCT) across 20 days, as recommended by the tool guidelines. |
| high baseline (resilient) | Staff roster set to meet 90<sup>th</sup> percentile of demand measured by the Safer Nursing Care Tool SNCT across 20 days. Designed to meet demand through permanent staff on 90% of days if all rostered staff attend. |
| Scenarios – variation in temporary staff availability |  |
| No temporary staff | No temporary staff available |
| Observed | Redeployments and empirical availability of temporary staff (<50% chance of fulfilled requests for all staff types and times of day). Based on data for temporary staffing request fulfilled for one of the participating hospitals see supplementary material, Table 5. |
| Higher availability | Redeployments and bank/agency staff requests each have 50% chance of being fulfilled. Thus this assumes higher availability of temporary staff than the core assumption. |
| Unlimited availability | Redeployments and bank staff requests each have 50% chance of being fulfilled and agency staff requests have 100% chance of being fulfilled. |

*\*All staffing plans make use of flexible deployment of float staff (from overstaffed units) and temporary staff hired from bank or agency when required and if available.*

#### Simulation Model

We developed a simulation in the software AnyLogic researcher edition version 8.3.2.(The AnyLogic Company, 2019). The simulation is an example of a Monte Carlo simulation since many input parameters (such as absence rates and demand for nurses) are stochastic, so are modelled as random variables following probability distributions. Figure 1 shows the main simulation steps, and both a video of the simulation in action (Saville and Monks, 2019) and a detailed model description following the STRESS reporting guidelines for agent-based simulations (Monks et al., 2019) are available (see Supplementary material Supplementary material.

**Figure 1.**
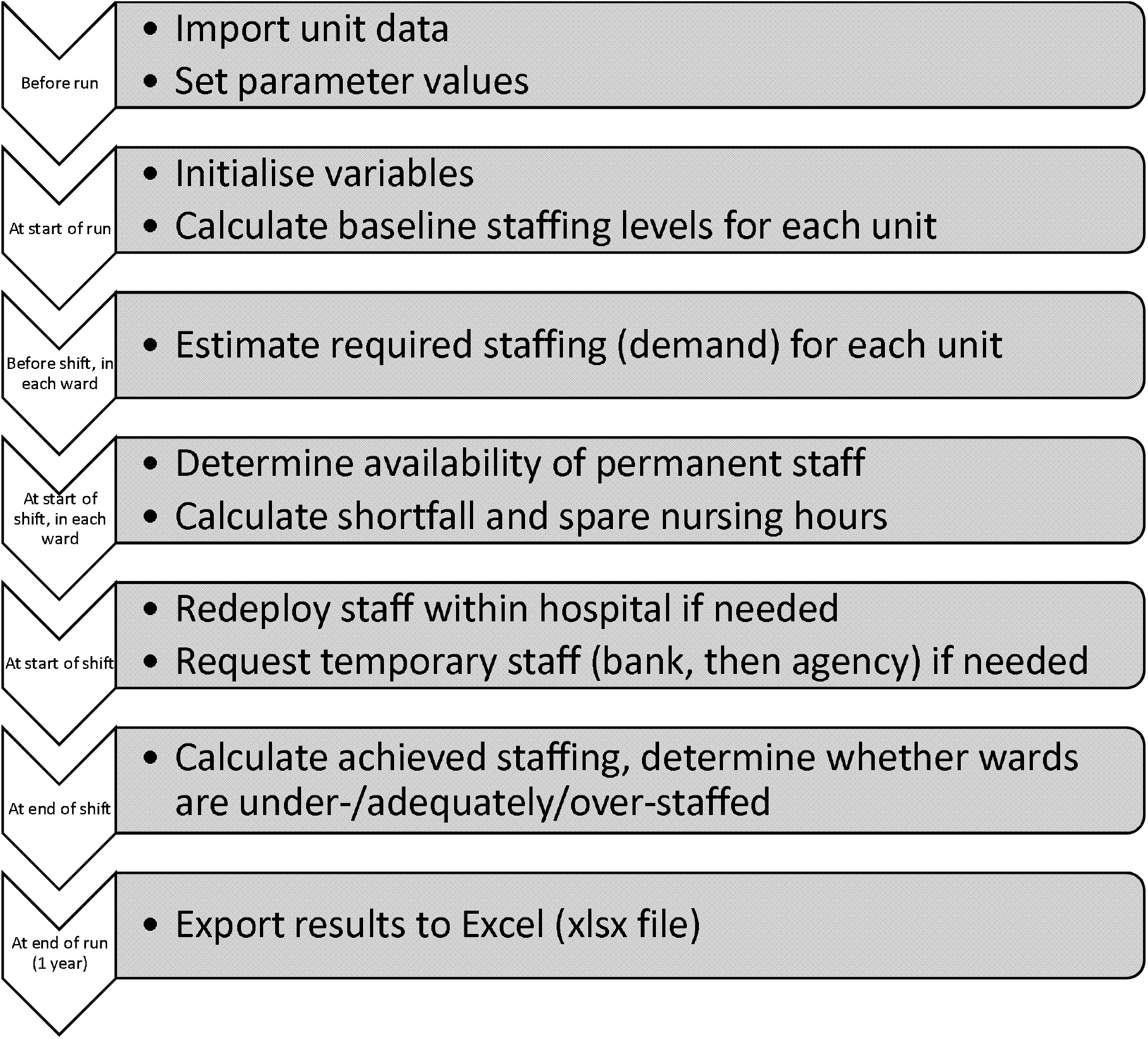
Flowchart of simulation steps adapted from(Griffiths et al., 2020b) with permission.

Appendix 1).

Daily demand for nursing care was simulated using the Safer Nursing Care Tool acuity dependency measure (The Shelford Group, 2014). To provide parameters for our models, we undertook an observational study over one year (2017) in 81 general (adult) medical/surgical inpatient units in three hospitals, in London, South East and South West England (Griffiths et al., 2020b, Saville et al., 2020a). Because estimates for mortality and the effect of low staffing on length of stay used in the present analysis (see below) applied to general medical and surgical populations, we excluded a specialist cancer hospital that participated in the parent study from this aspect of the study.

The base number of staff to be scheduled on each unit / shift was determined by taking a 20-day sample of daily patient acuity and dependency measures from each unit, as recommended when using the Safer Nursing Care Tool for establishment setting (The Shelford Group, 2014). The number of full-time nurses to be employed (the establishment) was converted into daily hours, removing the uplift for annual leave, staff education and other time away from the unit. The daily staff hours were distributed across shifts based on the distribution observed on each unit. Similarly the skill mix between registered nurses and assistants was based on the observed skill mix for each unit.

In the simulation, units are the ‘agents’, which move between being understaffed, adequately staffed or overstaffed as supply of staff and demand from patients varies. The daily census and the acuity/dependency of patients, varied based on parameters derived from the observational study, was used to simulate variable demand for nursing care. To reduce computational complexity, we did not model the year in order but rather took a random draw from the demand data for each ward for each shift. Where units had experienced substantial changes that might necessitate different numbers of staff scheduled, e.g. extra beds in winter, we only used the data from the unit before the change.

Additionally we included one-to-one specialing requirements in the demand measure (Wood et al., 2018), based on the observed use of such staff in each unit. Starting with a base number of staff rostered on each unit on each shift, the model simulates the short-notice staff absence due to sickness. The model then attempts to address any staffing shortfalls relative to that day’s demand from patients, firstly by redeploying (floating) staff from overstaffed to understaffed units, secondly by hiring staff from an internal bank of hospital employees and thirdly hiring from an external agency.

In all cases, registered nurses are substituted for registered nurses, and assistants are substituted for assistants. A threshold of 15%, corresponding to the tolerance used in the RAFAELA staffing tool, was used to trigger attempts to fill staff shortfalls (Fagerström et al., 2014). We assumed rates of unanticipated absence through short notice sickness of 4% for assistants and 3% for registered nurses, approximating known differences in sickness absence between these groups (Dall’Ora et al., 2019a). Current national sickness rates for all nursing staff are 4.5% (Moberly, 2018), but longer term sickness absence can be anticipated and temporary staff or overtime added to the roster and so this was not considered separately. The availability of temporary staff was, initially, based on the empirical availability to fill short notice requests as reported by one of the participating hospitals. Temporary staff availability varied by time of day and day of the week but was always less than 50%, often considerably so (see supplementary material Appendix 2, Table 5). Because such constraints are likely to be dependent on local labour market conditions, we also considered both a higher and unlimited availability of temporary staff as well as a scenario where there was no availability of temporary staff and the only flexible staff availability was floating staff from over staffed units (see Table 1). All allocations, hires and redeployments are subject to constraints about whole people, who must be deployed for a half/whole shift. For each shift, we calculated the achieved staffing relative to patient need.

### Model validation

Throughout model development, we performed verification (checking correct implementation of the model in simulation software) and validation (checking that we built an appropriate and sufficiently accurate model). Full details of the validation are given in Griffiths et al (2020b). We worked closely with nurses with responsibilities for workforce at the participating hospitals, who agreed assumptions and sense-checked results. We also presented and discussed early versions of the simulation model and results with the project steering group, which included nursing research, mathematical modelling and nursing workforce experts. We tested the model’s sensitivity to a number of key assumptions and found that neither staffing costs nor the rate of over / understaffed shifts appeared to be greatly affected by most assumptions such as the sickness / absence rates or the relative efficiency of temporary staff. The only assumption that made very large differences to the parameters estimated by the model related to the availability of temporary staff.

The estimated required staffing levels corresponded closely to the staffing actually deployed in the participating hospitals (Griffiths et al., 2020b). Estimated daily staff costs were similar (£140-150 per patient day) to actual costs. Since we assumed that hospitals were able to employ the staff needed to fill baseline rosters and did not consider staff rostered to cover longer term sickness, the level of temporary staff actually used in the participating hospitals was generally higher than in our models, as were the staff costs. We ran the model 10 times for each hospital and for a range of staffing scenarios, and calculated 95% confidence intervals around the means to assess the errors around the estimates. The confidence intervals were narrow, for example, for the standard staffing scenario, widths were <£0.25 cost per patient day.

### Staffing scenarios and cost effectiveness

We used the model to conduct a series of simulated experiments to explore the effect of different staffing plans on achieved staffing levels, the costs of staffing and patient outcomes by varying a number of parameters. The staffing plans and scenarios for temporary staffing availability are detailed in Table 1.

We took a limited perspective on costs, focussing on nurse staffing costs and reduced resources in terms of bed utilisation. The costs were for 2017 and no discounting was applied because of the short-term time horizon. Prices are in Great British Pounds (£) sterling. Using the 2017 Organisation of Economic Cooperation and Development (OECD) purchasing power parities, £1 had equivalent purchasing power to $1.46 US (https://data.oecd.org/conversion/purchasing-power-parities-ppp.htm). Hourly employment costs for substantive staff at each pay band were estimated using the mean costs for each band reported in the Unit Costs of Health and Social Care (Curtis and Burns, 2017). Salary and additional employer costs including pension contributions were included to derive a total cost for substantive and bank staff. Costs for agency staff were estimated using NHS guidance applicable to the study period, which set a cap on payment rates designed to reduce costs (NHS Improvement, 2018a). This therefore represents a low estimate. The daily unit staffing costs include allowances for working unsociable hours, in accordance with the Agenda for Change framework, using multipliers applicable to the study period (Nursing Times, 2008). See supplementary material Table 6.

The effects of each alternative staffing plan on length of stay and the risk of death relative to the standard plan were estimated using regression coefficients from a recent longitudinal study of the effect of variation in nurse staffing on patient outcomes (see supplementary material Table 7) undertaken in one of the participating hospitals (Griffiths et al., 2018, Griffiths et al., 2019). This study was chosen because of the direct connection to staffing data for the current study and the robust longitudinal design using patient level exposure to variation in nurse staffing as the independent variable. All alternative estimates we identified were from cross-sectional studies or different countries. Although direct comparisons with other studies of patient level exposure with longitudinal design is not possible due to different measures of low staffing, the observed effects were broadly similar in magnitude (e.g.Needleman et al., 2011).

We used data from the ‘standard’ staffing plan to determine a unit mean staffing level against which low staffing was judged. We summed the days of low staffing across all units. We calculated the mean change in staffing levels associated with each staffing plan. As there is some evidence that using high levels of temporary staff (>1.5 hours per patient day) can have an adverse effect on mortality (Dall’Ora et al., 2019b) we also summed days with high temporary staffing. From data for each unit on each day, we calculated the overall risk of exposure to low staffing and high temporary staffing. We estimated both staff costs and net costs after taking into account the value of bed days saved. The cost of changes in length of stay were estimated using the 2017/18 national average reference cost for a non-elective excess bed day (£337), a likely conservative assumption as it assumes that there are no specific treatments costs associated with extended stay.

We identified mortality rates and average length of stay for the hospitals from published data (NHS Digital, 2018b) and used these as the assumed baseline for the standard staffing plan. We calculated the change in the number of deaths by subtracting the number of deaths associated with the ‘standard’ staffing plan from the number of deaths estimated in the new scenario ((_Δ_exposure * Risk Ratio * baseline risk * population) – baseline death rate). We estimated ‘numbers needed to treat’ (NNT) or ‘numbers needed to harm’ (NNH) (Cook and Sackett, 1995) associated with each alternative plan using the formulae NNT=1/ARR or NNH=1 /ARI (where ARR and ARI are the absolute risk reduction and increase respectively). These figures represented the number of patients who would need to be exposed to a given staffing plan (on average) to ‘save’ (or lose) one additional life. We calculated the incremental cost effectiveness of alternative plans (_Δ_cost/_Δ_mortality) relative to the ‘standard’ plan. Our primary analysis considered the effect of low staffing only on mortality. Our secondary analysis additionally considered the effect of exposure to high levels of temporary staffing.

Given the narrow confidence intervals for parameters produced by the simulation (e.g. <£0.25 relative to mean daily costs of around £130) coupled with the computational time involved in running the models for multiple hospitals and multiple scenarios, data were generated from a single model run (365 days) for each of three different hospital configurations and we report results as an unweighted average of the three with incremental cost effectiveness ratios calculated as average change in outcomes / average change in costs. The underlying data generated by the model comprises 677,809 patient days and 29,565 unit days. We undertook sensitivity analyses to determine the impact of a number of parameters used in the economic model, including relative staff costs and the estimated effect of low staffing on mortality.

### Approvals and permissions

The study was prospectively registered (ISRCTN 12307968), ethical approval was granted by the University of Southampton (ergo ID 18809) and permission to undertake the research was granted by the Health Research Authority (IRAS ID 190548).

## Results

When availability of temporary staff was limited to the empirically observed level, the estimated staff cost per patient day for the standard staffing plan was £133, with a mean achieved staffing level of 3.6 registered nurse hours and 3.5 nursing assistant hours per patient day. Across all units achieved staffing was, on average, 9% below the measured requirement, with 90% of wards between 2% understaffed and 16% understaffed on average. Distributions of both required and achieved staffing varied by unit, but typically showed a slight positive skew (median 0.6). Sixteen per cent of units (13/81) showed a strong positive skew (>1) in required staffing and 14% (11/81) showed a strong positive skew in achieved staffing. Only one ward showed a strong negative skew in achieved staffing. Both required (median .7) and achieved staffing (median 1.1) tended to positive kurtosis, meaning there were more values in the tails of the distribution (further from the mean) than expected under a normal distribution, with 52% (42) of units having kurtosis of >1 for achieved staffing.

The achieved staffing level was lowest for the low baseline flexible staffing plan and highest for the high baseline resilient plan (Table 2) although differences between the plans reduced as the assumed availability of temporary staff increased. The achieved staffing levels and costs for the high baseline staffing plan were much less sensitive to changes in the availability of temporary staff than were the other plans.

**Table 2.**
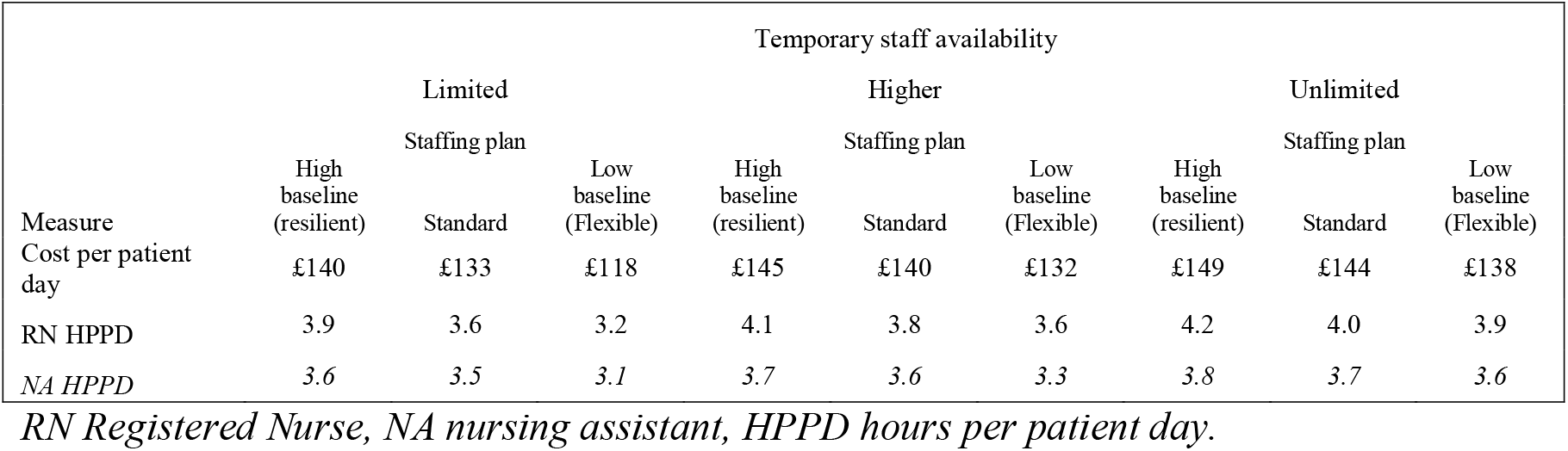
Achieved staffing levels and daily costs for different staffing plans with varying temporary staff availability.

Staffing plans with higher baseline staff (standard vs low baseline and high baseline vs standard) were associated with higher costs but shorter lengths of stay and fewer deaths (table 3). As the assumed availability of temporary staffing increased, differences between staffing plans reduced. Staffing plans with higher baseline staffing became more cost effective (less additional cost per improved outcome) as temporary staffing availability increased..

**Table 3.** Changes in Costs, effects and cost-effectiveness for standard vs low baseline (flexible) and high baseline (resilient) vs standard plans for varying levels of temporary staff availability.

|  | Staff cost |  | Bed days |  | Death |  | NNT (NNH) |  | Staff cost / life |  | Net cost / life |  |
| --- | --- | --- | --- | --- | --- | --- | --- | --- | --- | --- | --- | --- |
|  | Comparison |  | Comparison |  | Comparison |  | Comparison |  | Comparison |  | Comparison |  |
|  | High base (resilient) Vs Standard | Standard vs Low base (flexible) | High base (resilient) Vs Standard | Standard vs Low base (flexible) | High base (resilient) Vs Standard | Standard vs Low base (flexible) | High base (resilient) Vs Standard | Standard vs Low base (flexible) | High base (resilient) Vs Standard | Standard vs Low base (flexible) | High base (resilient) Vs Standard | Standard vs Low base (flexible) |
| **Low staffing effects only** |  |  |  |  |  |  |  |  |  |  |  |  |
| Temporary staff availability |  |  |  |  |  |  |  |  |  |  |  |  |
| None | 7.8% | 19.9% | -1.4% | -2.4% | -4.5% | -13.4% | 663 | 222 | £25,584 | £ 23,936 | £ 13,155 | £ 16,015 |
| Limited | 5.5% | 10.8% | -1.2% | -1.7% | -4.5% | -8.3% | 665 | 361 | £ 19,437 | £ 21,766 | £ 8,653 | £ 21,766 |
| Higher | 4.0% | 5.5% | -1.0% | -0.8% | -3.8% | -4.4% | 873 | 719 | £ 17,230 | £ 20,422 | £ 6,451 | £ 12,722 |
| Unlimited<sup>3</sup> | 2.9% | 1.6% | -0.9% | -0.3% | -3.0% | -1.9% | 1272 | 2828 | £ 15,612 | £ 10,141 | £ 3,693 | £ 4,520 |
| **Including temporary staffing effects** |  |  |  |  |  |  |  |  |  |  |  |  |
| Temporary staff availability |  |  |  |  |  |  |  |  |  |  |  |  |
| None | 7.8% | 19.9% | -1.4% | -2.4% | -4.7% | -14.6% | 632 | 204 | £ 24,364 | £ 22,013 | £ 12,528 | £ 14,729 |
| Limited | 5.5% | 10.8% | -1.2% | -1.7% | -4.7% | -10.2% | 627 | 296 | £ 18,404 | £ 17,537 | £ 8,193 | £ 10,568 |
| Higher | 4.0% | 5.5% | -1.0% | -0.8% | -4.4% | -11.1% | 703 | 305 | £ 15,067 | £ 7,623 | £ 5,642 | £ 4,749 |
| Unlimited<sup>3</sup> | 2.9% | 1.6% | -0.9% | -0.3% | -4.1% | -15.1% | 766 | 283 | £ 11,356 | £ 1,156 | £ 2,687 | £ 515 |

Where the availability of temporary staff was limited, the high baseline resilient staffing plan increased staffing costs by 5.5% whereas the low baseline flexible staffing plan reduced staff costs by 10.8% compared to standard plans. When the availability of temporary staff was unlimited, the high baseline plan was associated with a 2.9% increase in staff costs while the low baseline plan was associated with a reduction of 1.6%.

With limited temporary staff availability, the high baseline resilient staffing plan was associated with a 1.2% reduction in the average length of hospital stay, and a 4.5% reduction in the relative risk of death, equating to one life saved for every 665 patients admitted to a hospital (number needed to treat). By contrast, with the low baseline flexible plan, many shifts were left critically understaffed, and consequently there was a 1.7% increase in the average length of hospital stay and an 8.3% increase in the risk of death, equating to one additional death for every 361 patients.

The outcomes for the low baseline flexible staffing plan were more sensitive to the availability of temporary staff than were those for the resilient plan. If no temporary staff were available, then deaths were increased by 13.4% with a flexible staffing plan (relative to the standard plan), whereas with unlimited temporary staff availability deaths are increased by only 1.9%. For the resilient plan the equivalent range was a reduction in death ranging from 4.5% with no temporary staff availability to 3% with unlimited temporary staff.

Compared to the low baseline flexible plan the standard staffing plan staff cost per life saved were £ 21,766 when availability of temporary staff was limited. Much of the additional staff cost is offset by the value of reduced hospital stays and so the net cost per life saved was £ 13,117. Similarly, compared to the standard plan, staff costs per life saved associated with the higher baseline resilient plan were £19,437. More than 50% of this cost was offset by the value of the reduced length of stay, leading to a net cost of £9,506 per life saved.

Although the adverse effects of plans with lower baseline staffing were reduced with higher temporary staff availability, the relative cost-effectiveness of higher staffing was more favourable under these circumstances. For example, with unlimited temporary staff availability the net cost per life saved for the standard plan relative to the flexible plan is only £4,250, while the cost per life saved for the resilient plan relative to the flexible plan was reduced to £3,963.

Our primary analysis assumed that temporary staff are as effective as permanent staff and so adverse outcomes result from understaffing alone. There is some evidence that high levels of temporary staffing can have an adverse effect on patient outcomes and so we also considered these additional adverse effects of temporary staffing on mortality (see table 3). The estimated mortality associated with lower baseline staffing was increased when considering an adverse effect from high levels of temporary staff. In the primary analysis higher availability of temporary staff tended to reduce the difference between plans and mitigate the adverse effects of lower baseline staffing. This was not the case when an adverse effect of high temporary staff was included. When unlimited temporary staff were available the low baseline flexible staffing plan was associated with a 15.1% increase in mortality compared to the standard plan.

Consequently, if a negative effect from high temporary staff was assumed, the cost effectiveness of higher baseline staffing was further improved, particularly when comparing standard staffing to the low baseline ‘flexible’ staffing plan and when temporary staff availability was higher. For example, the net cost per life saved for the standard plan compared the low baseline plan was only £515 per life saved (compared to £4250).

### Sensitivity analyses

We undertook a series of sensitivity analyses. Although the pattern of results was largely unchanged by variation in model parameters, the magnitude of differences between staffing plans was sensitive to core parameters in the model, although these differences were generally unlikely to change substantive conclusions about cost effectiveness. Table 4 illustrates this by showing the change in net cost per life saved for the high baseline resilient staffing plan relative to the standard plan for limited and unlimited availability of temporary staff associated with alteration of some core parameters.

**Table 4.** Effect of changing model parameters on net cost per life saved for high baseline resilient vs standard staffing plan with limited and unlimited availability of temporary staff.

| Parameter alteration | Change in cost per life saved estimate |  |
| --- | --- | --- |
|  | Limited temporary staff | Unlimited temporary staff |
| Mortality estimate at upper 95% CI | -£3,705 | -£2,187 |
| Mortality estimate at lower 95% CI | £18,377 | £11,019 |
| Additional bed day cost + 25% | -£2,729 | -£2,736 |
| Cost of agency staff + 25% | -£870 | -£4,312 |
| Cost of agency staff=cost of bank staff | £348 | £1,724 |
| Cost of bank staff=cost of permanent staff | -£233 | -£1,105 |
| No floating of staff between units | £2,477 | £2,146 |
| All costs increased by 25% | £2,377 | £1,380 |

The most significant sensitivity was the estimated effect of low staffing on mortality. Taking the upper bound of the 95% confidence interval for the mortality effect considerably reduced the cost per life saved, whereas taking the lower bound increased it. With limited availability of temporary staff net cost per life saved was increased by £18,377 if the low bound estimate of the effect on mortality was used.

In our original model, we assumed that bank staff were cheaper than permanent staff (because pension costs were reduced) and that agency staff were paid at the rate capped by NHS Improvement guidance. Changing these assumptions, including changing the assumptions so that bank and agency staff had similar costs (equivalent to sourcing all temporary staff from the bank) made most difference when unlimited availability of temporary staff was assumed and only small differences when availability was limited. NHS. A 25% increase in agency staff costs substantially reduced the incremental cost per life saved when temporary staff availability was unlimited.

If no floating of staff from overstaffed units to understaffed staffed units was permitted, the cost per life saved associated with the resilient staffing plan increased, although the magnitude of the difference was small relative to the impact of assumptions about the availability of temporary staff (see supplementary material Table 8).

The main model results are based on the unweighted average of models built with data and unit configurations of three different hospitals. Individual hospital results varied in their magnitude but the pattern of results for the relative costs or effects of different staffing plans and the impact of varying availability of temporary staff was generally consistent (supplementary material Table 9). However, for one hospital, the staff costs under the flexible staffing plan were marginally more expensive than for the standard plan if there was unlimited availability of temporary staff. In all cases the net cost per life saved for the resilient staffing plan was <£15,000 when temporary staffing was available.

## Discussion

In this study, we undertook a series of simulation experiments to understand the relative cost-effectiveness of three staffing plans. The plans differed in the level of baseline nurse staffing planned on the roster (schedule) but all used flexible deployments to meet daily variation in demand. Although low baseline staffing and use of flexible staff deployment to meet peaks in demand is often assumed to be economically efficient, we found that staffing plans with higher baseline staffing appeared to be cost-effective compared to plans with lower baseline staffing.

Under a staffing plan with baseline staffing set at a low level, where the use of flexible staff to meet varying need was emphasised, we found that variation in demand could not be effectively addressed by floating staff from overstaffed units or hiring temporary staff whose availability is limited. Although staff costs were reduced when compared to a standard plan, set to match (mean) average demand, the apparent economic efficiency was achieved in large part because many shifts were left understaffed, leading to adverse consequences for patient care. Patient deaths were increased. Because length of stay was also increased the net cost savings were much less than the apparent saving on staff costs indicated. As the availability of temporary staff increased, understaffing was reduced but so were the cost savings compared to the standard staffing plan. Conversely, a higher baseline staffing plan, which emphasised resilience and was set to meet most peaks in demand, was more expensive but delivered better outcomes. Additional costs were partly offset by savings from reduced hospital stays.

In the face of ongoing nurse shortages, a flexible staffing policy that relies on a minimal core staff and makes heavy use of temporary assignments to deploy scarce staff to meet need seems highly appealing. The priority is, by implication, to avoid the apparent inefficiency of ‘overstaffing’, treating it as a risk to be avoided that is equivalent to understaffing (Saville et al., 2019). However, our findings challenge a number of key assumptions underlying such an approach. Distributions of demand are not symmetrical for many units, and understaffing was far more common than overstaffing in both our simulation and our empirical observations (Griffiths et al 2020, Saville et al., 2020b). With a low baseline, the risk of understaffing is higher and there is less likelihood that any given unit is overstaffed. Therefore fewer staff are available to float to understaffed units. Consequently, the demand for temporary staff from other sources (in this study an internal bank or an external agency) is increased, as is the risk that shifts remain unfilled.

Any strategy designed to balance the risk of overstaffing and understaffing must recognise that there is considerable ‘salvage value’ associated with the apparent excess of staff (Davis et al., 2014a). Any excess staff clearly have value because they could be deployed elsewhere, but when not redeployed they will still be adding value directly on their home unit even though apparently ‘surplus’ to requirements. This is because the criteria used to identify the required staffing level is not set at a level beyond which there is zero or a diminishing effect from adding additional staff. Instead, the target staffing level is defined to meet a subjectively defined level of quality (Fagerström et al., 2014, The Shelford Group, 2014) and there is evidence that outcomes and quality of care improve further when staffing is at levels above those prescribed by current norms (Fagerstrom et al., 2018, Griffiths et al., 2020b, Junttila et al., 2016). Furthermore, higher nurse staffing is likely to contribute more value beyond that considered here, where we focus on a single effect, the risk of death. There are other benefits associated with higher staffing that are of value to patients, including avoidance of adverse events that fall short of death and improved patient experience (e.g.Bridges et al., 2019, Shang et al., 2019).

With many countries experiencing staff shortages, the goal of higher baseline staffing may seem simply unachievable. However, it is only when the availability of temporary staff is not limited at all that the low baseline flexible staffing plan appears to be viable in terms of avoiding a substantial negative impact on patient outcomes. Under these circumstances, the cost savings are largely eroded and the cost per life saved associated with staffing plans using higher baseline staff deployed on units is modest. Furthermore, if adverse effects from high levels of temporary staff are considered, adverse outcomes could still remain at a high level. Perhaps most striking of all in the context of labour market shortages, under these circumstances the average achieved staffing levels under the flexible staffing policy come close to those of the other policies. In our models, we assumed that temporary staff were equally as efficient as permanent staff. In some cases, for example where the flexible staff are in fact staff from the home unit undertaking overtime, this assumption may be warranted but overall this is unlikely to be the case (Duffield et al., 2020). There is little indication that this flexible staffing plan achieves a more efficient deployment of a scarce staffing resource or substantially reduces overall demand for that scarce resource. Rather it may simply be an inefficient way of deploying much the same resource.

Superficially, our findings appear to contradict those of Kortbeek et al. (2015) and others who have modelled the benefits of flexible staffing. However, many previous modelling studies have been based on achieving particular nurse to bed or patient ratios without taking into account variation in need at the patient level or the impact on quality and outcomes (Saville et al., 2019). Kortbeek et al described a flexible staffing model based on hourly bed census predictions as ‘efficient’, although the productivity gains relative to a fixed staffing solution were modest for most of the scenarios considered (Kortbeek et al., 2015). In our study all the scenarios considered were ‘flexible’ in so far as they all call on redeployments and temporary staff to meet varying demand. Our study focussed on the best approach to determining the baseline staff levels in the face variation and real life constraints on staff availability. Similar to Kortbeek et al. (2015) we show that floating of staff between units makes a modest contribution and appears to improve the cost-effectiveness of staffing plans with higher baseline staffing levels relative to lower. Crucially, neither Kortbeek’s result or ours can be used to support low baseline staffing levels as a means to achieve an efficient flexible staffing policy.

Even if the only consideration was reduced staff costs, savings from lower baselines are smaller than assumed because of increased hospital stays, which result from complications and delays in discharge preparation (Needleman et al., 2006). Little saving is made when there is unlimited temporary staff availability, but outcomes remain worse in part because of the ‘trigger effect’ whereby short staffing has to be of sufficient magnitude to warrant an additional staff member being scheduled for a significant proportion of a whole shift.

Our findings are more consistent with those of Harper et al. (2010), who modelled demand for nursing care with a similar measure to that used in the current study. They concluded that in order to minimize costs, a hospital should employ more nurses than the average need would indicate. We found that although a staffing plan with higher base staffing incurred additional staff costs, much of the increased cost was offset by savings from reduced hospital stays and such plans are potentially cost-effective. Differences may arise because Harper et al. (2010) considered only permanent and agency staff, whereas our models assumed that the first call would be on floating staff and then staff from the internal bank, who are potentially cheaper than permanent staff because of reduced contributions to pensions for what is, in effect, overtime. Furthermore, Harper et al.’s study preceded attempts to constrain what were regarded as excessive charges to the National Health Service for such staff in the UK (NHS Improvement, 2018a). Perhaps most significantly, Harper’s study calculated the full cost of meeting need whereas our model included real world constraints, which meant that some shifts were understaffed. Understaffing associated with lower base staffing rosters accounts for most of the differences in staff costs between the staffing plans.

At the outset, we described a staffing plan with a higher baseline establishment as ‘resilient’ because it was designed to ensure that enough staff were rostered to meet predictable peaks in demand. A key characteristic of a resilient system is its ability to cope with stressors (Berg et al., 2018). In this respect our findings show that the resilient staffing plan does indeed cope with the ‘stress’ of varying demand more successfully than the alternatives. While our findings do not support the use of a flexible staffing plan with a low baseline staff roster, all our plans make use of flexible staffing to some extent. It is clear that some degree of flexibility has the potential to benefit patients and is likely to be superior to a fixed staffing plan and so flexible staffing does contribute to resilience.

Floating of staff between units in our simulation made a positive contribution to cost effectiveness. Other studies have highlighted that modest use of internal redeployments have the potential to be beneficial, although unconditional use of floating staff between units is problematic and may lead to poor outcomes (Maenhout and Vanhoucke, 2013). The presence of staff who are unfamiliar with the unit means that such staff are unlikely to be as efficient as permanently assigned staff (Duffield et al., 2020). The potential harms associated with use of temporary staff might be minimised by the use of dedicated float pools of staff who are specifically trained to work in a range of areas and rostered with the intention of being available to float between units, although there is limited direct evidence (Dall’ora and Griffiths, 2018). Assuming that such float pools are themselves routinely rostered there is still a need to determine the home unit on which a staff member is deployed unless floated. The resilient staffing plan modelled here could provide the basis of such a float team system, with some staff from the float team rostered on to units to deal with the anticipated peaks in demand but available to be floated to other units if conditions permitted and required it. Depending on the overall level of demand, baseline rosters under such a system might be even higher, reducing the requirement for additional temporary hires.

Unless very pessimistic assumptions about life expectancy or utility of life gained were made for our estimates, it seems likely that the cost-effectiveness estimates for the cost per life saved for the resilient staffing plan would lead to costs per quality adjusted life year that sit well below generally accepted thresholds for defining cost-effectiveness (Marseille et al., 2014). An intervention that costs less that the annual per capita gross domestic product per disability adjusted life year is regarded as highly cost effective (Marseille et al., 2014). The UK per capita gross domestic product in 2017 was £39,977. If we assume that each death averted in the current model achieves at least one disability adjusted life year, higher baseline staffing was always cost effective, even under the most pessimistic assumptions about the effect of higher staffing on mortality.

Other more stringent thresholds have been suggested for cost effectiveness have been proposed (Claxton et al., 2015) and the National Institute for Health and Care Excellence, the independent body charged with evaluating costs and effectiveness for treatments to be provided by the publically funded universal National Health Service, categorises drugs that cost £10,000 per quality adjusted life year as providing ‘exceptional value for money’. Depending on the achieved life expectancy it is possible that higher baseline staffing meets even these more stringent thresholds for cost effectiveness. Additionally, if the potential adverse effects of temporary staff are considered, the economic argument for the higher baseline resilient staffing plan, with its lower use of temporary staff, becomes more compelling.

On the other hand, because the ‘flexible’ staffing plan is associated with worse outcomes than the standard, it is easy to reject it simply because it is not an effective strategy. Given that the standard approach to determining baseline staff is the norm, the relative cost-effectiveness does not support a case for disinvestment and a move to a lower baseline. Under circumstances where the low baseline flexible staffing plan comes closest to providing an acceptable alternative, the relative cost-effectiveness of the standard staffing plan approaches the level at which the standard plan would dominate in terms of economic decision making, because the cost per life saved associated with it is so low. Indeed, in one of our hospital models the standard plan proved to be both cheaper and more effective.

## Limitations

The use of simulation allows experiments about different staffing configurations on a scale that would be simply unfeasible in real life. However, although our results appear to be robust to variation in many of the assumptions in our model, and the simulation was extensively validated, the results are, nonetheless simulated. We considered only a limited range of costs and it is possible that adverse events associated with low staffing generate additional costs for the hospital, which would tend to further reduce the cost per life saved for staffing plans with higher baseline rosters. The pattern of results was consistent across the three hospitals in the model but the magnitude of the differences between staffing plans is sensitive to the hospital configurations and so cannot be generalised to other hospitals. The estimates of the effects of nurse staffing came from a robust longitudinal study undertaken in one of the hospitals that provided data for the models. While the evidence that nurse staffing plays a causal role in variation in patient outcomes seems compelling, unmeasured confounding and other shortcomings nonetheless potentially bias the results of observational studies. However, the substantive conclusions about the relative benefits and cost-effectiveness of higher baseline staffing were not altered when assuming much lower adverse effects of low staffing.

We did not directly model any efficiency or effectiveness loss associated with use of temporary staff, although there is some evidence that suggests that there is some loss of productivity and / or quality when temporary staff are deployed (Dall’ora et al 2018). Our secondary analysis considered the potential adverse outcomes associated with heavy use of temporary staff producing results that were substantially more favourable for higher baseline staffing levels.

The underlying data on bank staff used to develop the models made no distinction between staff who were working voluntary overtime on their own unit and other staff employed through the bank, who would be less familiar with the host unit. Therefore we were unable to account for differences in these approaches to flexible staffing in our model. The adverse effect of high levels of temporary staffing we modelled may be somewhat sensitive to the mix of staff, although use of overtime has also been associated with adverse outcomes. However our results are not substantially altered either this effect or the relative costs of staff groups. This limitation seems unlikely to change substantive conclusions.

As our model did not seek to predict patterns of demand, rather to estimate the average effect of a fixed baseline staffing level, we sampled data for each day at random and did not consider serial correlations. This simulates the average effect over a period with a fixed permanent roster with short term responses to varying demand by deploying temporary staff. If typical demand levels changed in the medium to long term, for example due to a seasonal pattern, baseline staff levels would need to be altered accordingly. Thus our model is consistent with guidance from the National Institute for Health and Care Excellence in England, which recommends a review of unit staffing plans at least twice a year, with additional reviews when changes such as patient case mix, which could alter demand, occur (National Institute for Health and Care Excellence (NICE), 2014).

## Conclusions

Flexible staffing plans that attempt to make the best use of a scarce nurse staffing resource by minimising the staff that are routinely rostered are likely to harm patients because temporary staff may not be available at short notice. Such plans are not efficient or effective solutions to nurse shortages. When sufficient temporary staff are available, there is little reduction in costs or the overall number of staff required, and so this approach does little to address nursing shortages.

A staffing plan using flexible deployments with a low number of staff on the baseline roster is not resilient, because it is unable to properly meet varying demand. In contrast, a plan with a higher baseline staffing set to meet predictable peaks in demand is both resilient and more flexible, because the ‘excess’ staff are productive. In the context of a sufficient baseline establishment, flexible staffing, including floating staff between units, can be guided by shift-by-shift measurement of patient demand, but proper attention must be given to ensuring that the baseline number of staff rostered is sufficient to meet at least average demand or higher. The apparent risk of overstaffing is unlikely to materialise because the additional staff contribute to flexible staffing when available to be floated between units and enhance quality and safety if they remain in the home unit. Staffing plans with higher baseline staff levels are highly cost effective.

## Data Availability

This paper draws on research and data reported in more detail in the NIHR Journals Library Health Services and Delivery Research. The data for this paper consist of anonymous ward and hospital parameters and simulation results. All data requests should be submitted to the corresponding author for consideration. Access to available anonymised data may be granted following review. The simulation model and accompanying documentation are available from the author on reasonable request.

https://doi.org/10.3310/hsdr08160

## Acknowledgements

*The Safer Nursing Care Team study team comprise:* Griffiths, Peter^1,2,3^ *; Saville, Christina^1,2^; Ball, Jane E^1,2^; Jones, Jeremy^1^; Monks, Thomas^4;^ Chable, Rosemary^5^; Dimech, Andrew^6^; Jeffrey, Yvonne^7^; Mauruotti, Antonello^8^; Pattison, Natalie^9,10^; Recio Saucedo, Alejandra^1^; Sinden, Nicola^3^

Additionally, Aspden, Clare^5^, Cassar, Tracey^3^ and Hunter, Shirley^*7*^ contributed to data collection and Lambert, Francesca contributed project management and advised on patient and public involvement.

^1^ University of Southampton, UK

^2^ National Institute for Health Research Applied Research Centre (Wessex), Southampton, UK

^3^ Portsmouth Hospitals University NHS Trust, UK

^4^ University of Exeter, UK

^5^University Hospital Southampton NHS Foundation Trust, Southampton, UK

^6^The Royal Marsden NHS Foundation Trust, London, UK

^7^Poole Hospital NHS Foundation Trust, Poole, UK

^8.^ Libera Università Maria SS. Assunta, Rome, Italy

^9^ University of Hertfordshire, UK

^10^ East and North Herts University NHS Trust, UK

An extended account of this study was published as part of the NIHR Journals Library as: Griffiths, P., Saville, C., Ball, J.E., Chable, R., Dimech, A., Jones, J., Jeffrey, Y., Pattison, N., Saucedo, A.R., Sinden, N., Monks, T., 2020. The Safer Nursing Care Tool as a guide to nurse staffing requirements on hospital wards: observational and modelling study. Health Serv Deliv Res 8, 16.10.3310/hsdr08160

## Supplementary material

### Appendix 1 Simulation model technical details

*Reproduced with permission from: Appendix 1* Griffiths, P., Saville, C., Ball, J.E., Chable, R., Dimech, A., Jones, J., Jeffrey, Y., Pattison, N., Saucedo, A.R., Sinden, N., Monks, T., 2020. The Safer Nursing Care Tool as a guide to nurse staffing requirements on hospital wards: observational and modelling study. Health Serv Deliv Res 8, 16. (Changes to original: abbreviations spelled out and cross-references amended.)

#### Implementation and experimentation

The simulation was developed on a Windows 10 Enterprise operating system version 1709, build number 16299.726 in Anylogic 8 researcher edition software version 8.3.2, build number 8.3.2.201807061745 x64. This used Java 2 Standard Edition 8.0 and Fluid library version 2.0.0.

We used the default random number generator in Anylogic: an instance of the Java class Random, which is a Linear Congruential Generator (LCG).

Time is modelled as fixed time steps. There are no concurrent events that are interdependent.

There is no warm-up period or model initialisation since each day/shift is independent of the next. The model is stochastic. The run length is one year, and the time units are six-hour shifts (the model also works for days). The number of replications is 10.

Running one experiment (1 year, 10 runs) takes between 10 and 20 minutes for the different hospital Trusts on a Dell Latitude laptop with Intel® Core™ i5-7300U processor with 2.60GHz CPU speed, 8 GB RAM and 32 GB ROM.

#### Computer Model Sharing Statement

For a video demonstration see https://eprints.soton.ac.uk/430632/. The simulation model is available from the corresponding author on reasonable request. Anylogic simulation software can be downloaded from https://www.anylogic.com/downloads/.

#### Simulation model logic

An overview of the simulation logic is shown in FIGURE 2. Each stage is described in more detail below. The simulation was designed to work for any number of wards. The wards are ‘agents’ which interact with each other (within directorates) by lending/borrowing staff in an attempt to cover shortfalls of staffing in one ward with surplus in another. Outputs are updated at the end of each time step (day/shift) and aggregated at the end of each year.

**FIGURE 2.**
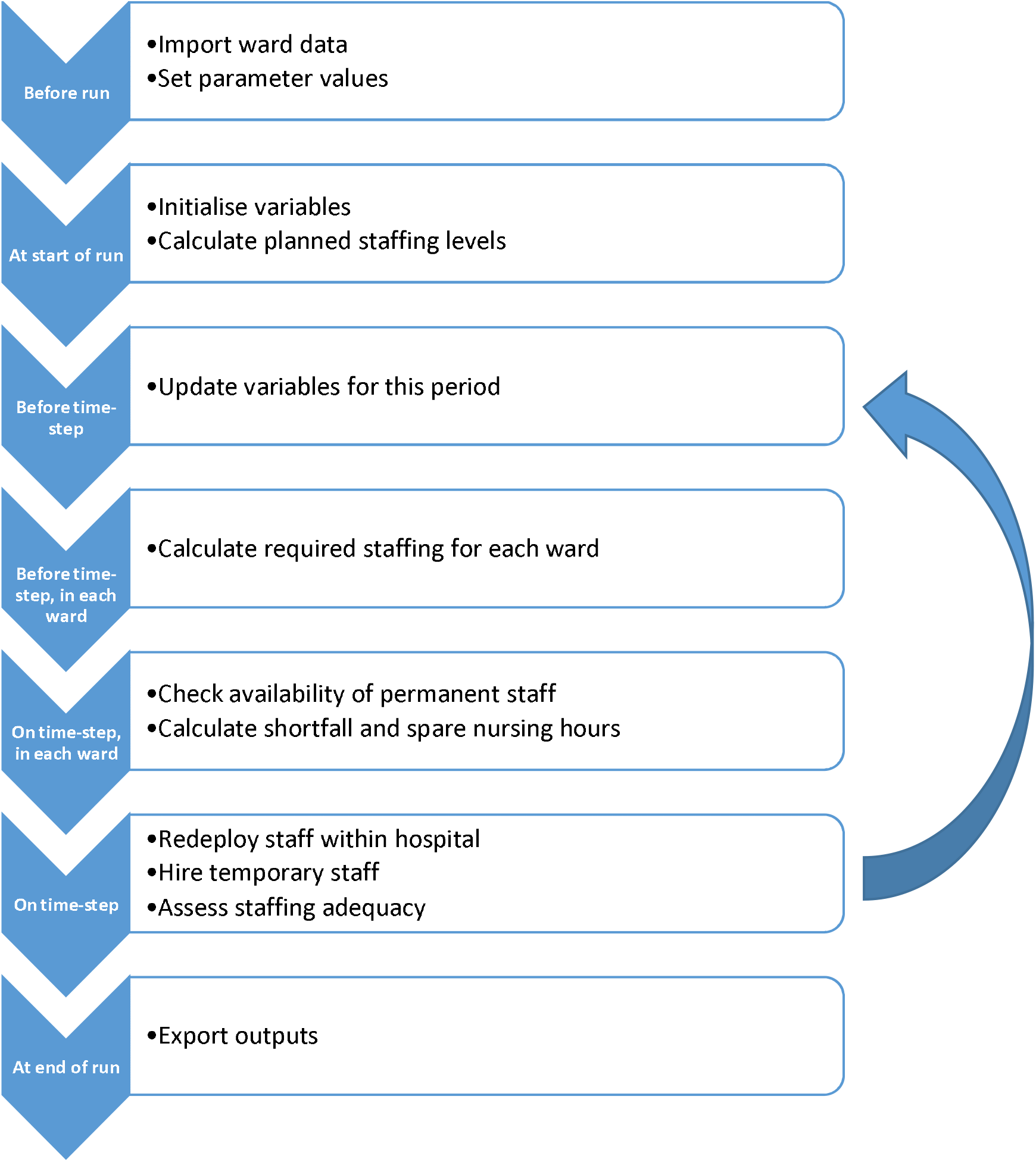
Simulation model overview diagram.

##### Before running the simulation

Before running the simulation for a particular hospital Trust, information about its wards is imported from Excel files to fill database template tables within the simulation. The three files contain occupancy data, acuity/dependency data and other ward information. The occupancy distributions consist of the number of patients, labelled with the ward identification numbers (a contiguous number for internal purposes and the number used for external reporting), day of the week and 6-hour-shift. The acuity/dependency distributions consist of the proportions of patients in each SNCT acuity/dependency level and the proportion of patients that require specialing, labelled with the ward identification numbers and observation number (used when sampling a row). The other ward information consists of the baseline staffing levels for each scenario (in WTE), the skill mix (in the morning, afternoon, evening, night, and over a 24-hour period), the distribution of staff over 24 hours (proportion of staff deployed in the morning/afternoon/evening/night), the directorate number, whether the ward is an acute admissions unit and some purely descriptive information (directorate, medical or surgical).

The probabilities of requests for temporary staff being fulfilled is imported from an Excel file into another database template table. The template allows probabilities to differ between bank and agency, up to three staff types, weekend and weekday, and time period (morning, afternoon, evening, night, and over a 24-hour period).

Next, further parameter values that apply globally to the whole hospital Trust (the system) are set. These parameters relate to:

1. nursing staff requirements - acuity/dependency multipliers, specials multiplier, standard deviation of multipliers, minimum number of registered nurses needed (constraint), demand for registered nurses with a particular skill
2. permanent staff - the baseline staffing level, the data sample used to calculate the staffing level, the staff types, absence chance for each staff type, absence length, proportion of registered nurses with a particular skill
3. redeployed staff - redeployment rules, priority sequences for providing and receiving redeployed staff, efficiency of redeployed staff, redeployed staff shift length
4. temporary staff - rules for requesting temporary staff, efficiency of bank and agency staff, temporary staffshift length
5. display settings - the understaffing criterion to plot in charts, e.g. 15% or more under requirement
6. eneral settings - step length, round down bound used when converting required hours to requested hours.

##### At start of run

At the start of a run, variables tracking time (the step number and the shift number) are reset to zero. The wards, staff types and sharing groups are counted. Occupancy distributions are created from the occupancy data table. Data are placed in arrays, which are convenient structures for working with multi-dimensional data.

Then, the establishment (number of staff employed in WTE) is converted to the number of planned deployed hours per time step (6-hour-shift or day), including applying the skill mix, rounding to whole people and dealing with minimum constraints, as follows. Note that the establishment does not need to be a whole number since staff may work part-time, but the planned number of staff to deploy each six-hour shift should be a whole number. As in our other analyses we use equation 3 for converting the planned staffing in WTE to the planned total care hours per day (see Equation 3, Appendix 1, main report (Griffiths et al., 2020b).

The planned skill mix (proportion of staff that are registered nurses) and distribution of staff over the day in each ward is set as the average observed for that ward. There is a constraint that there must be at least one registered nurse present on each ward, so if the registered nurse hours is under 6, this is rounded up to 6. Otherwise, the registered nurse hours are rounded up or down to the nearest six hours. The remaining planned hours are assigned to nursing support workers, and again rounded up or down to the nearest 6 hours.

For example, suppose the planned nursing hours for a morning shift on a particular ward are 18, and the skill mix is 50%. This is equivalent to 9 hours of registered nurse time, which is rounded up to 12 hours. There are 6 hours left to cover which are assigned to nursing support workers.

The planned deployed hours per day (sum over the four shifts) are converted back into WTE to enable calculation of the cost of employing this number of permanent staff.

##### Before time-step

Before each time-step, i.e. before the simulation switches to the next period (six-hour shift or day), the variables for this period are updated. These variables are the time step, the shift (1 to 4), the day type (weekday, Saturday of Sunday/bank holiday) and the planned staffing level for this shift. The deployment array (numbers of staff from each source and of each staff type deployed on each ward in what capacity) is reset at zero ready to be filled in the next stages.

##### Before time-step, in each ward

Next, the required staffing for this period is calculated for each ward in turn. For this, firstly the number of patients on the ward is sampled from the occupancy distribution for that ward, day of week and shift (morning, afternoon, evening or night).

Secondly, the acuity/dependency profile is sampled from the acuity/dependency data. This is done by selecting a random observation for that ward (we assumed there were no day of week or time of day patterns). For each patient, the probability of being in each acuity/dependency category and the probability of requiring specialing are the corresponding observed proportions. The required staffing per patient (in WTE) is sampled based on the patient’s acuity/dependency category and specialing requirements. This is converted into the required staffing level for this period using the skill mix, distribution of staff over the day and minimum constraints (as for the planned staffing levels), but is not rounded.

##### On time-step, in each ward

On the time-step, i.e. immediately when the period begins, the number of hours of staffing provided by permanent staff in this period is calculated for each ward. For this, the number of planned staff who are not unexpectedly absent (i.e. at short notice) is calculated. The chance of being unexpectedly absent can differ between staff types in the model. All these staff are allocated to their home ward to start with. The (absolute) shortfall for each staff type is calculated as required minus allocated hours. Where applicable, the simulation checks which of the registered nurses working are IV-trained (sampled probabilistically).

Similarly the spare hours (hours that could be redeployed to another ward) for each staff type is calculated. This is the allocated minus the required hours, rounded down to the nearest multiple of ‘redeployed hours chunk’, since staff can only be redeployed for fixed time periods.

##### On time-step

Next, staff are redeployed within sharing groups (directorates) to attempt to cover shortfalls for each staff type, as shown in FIGURE 3. The shortfall is rounded up or down (depending on the round down bound) to the nearest multiple of ‘redeployment chunks’. Requests for extra staff are triggered if the rounded shortfall for that staff type is more than zero, and if either the total shortfall or the staff type shortfall are more than the trigger (6 hours). In order to decide the priority of redeploying extra staff to wards, lists of wards are sorted using the bubble sort algorithm(Knuth, 1981), which works by comparing the shortfall as a proportion of the requirement (or spare hours as a proportion of requirement) for adjacent wards in the list and swapping them if they are in the wrong order.

**FIGURE 3.**
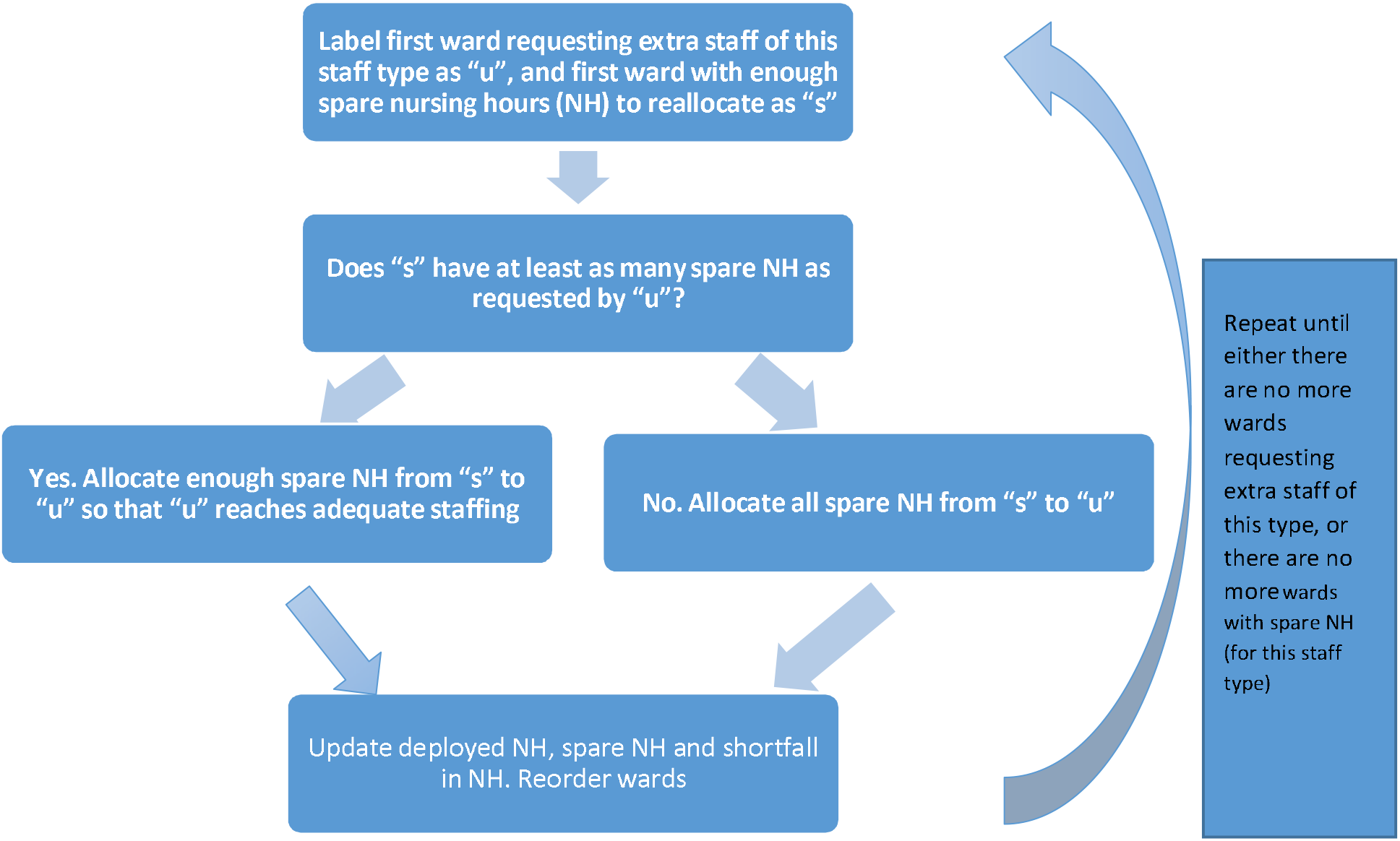
Summary process flow of staff redeployment.

Following this, for the wards that are still requesting extra staff, for each staff type, first bank and then agency staff are requested, as shown in Fig 3 above. The hours requested are the shortfall rounded up or down (depending on the round down bound) to the nearest multiple of ‘external work time’. The probability of a request for temporary staff being fulfilled depends on the staff source, staff type, whether it is a weekday or weekend and the time period.

### Appendix 2 Additional Tables

**Table 5.** Observed percentage of same-day and previous-day temporary staff requests filled.

| REQUESTS FULFILLED BY BANK STAFF | REGISTERED NURSES | NURSING ASSISTANTS |
| --- | --- | --- |
| **WEEKDAY** |  |  |
| MORNING | 28% | 28% |
| AFTERNOON | 10% | 9% |
| EVENING/NIGHT | 19% | 37% |
| **WEEKEND** |  |  |
| MORNING | 32% | 31% |
| AFTERNOON | 13% | 17% |
| EVENING/NIGHT | 19% | 32% |
| **REQUESTS FULFILLED BY AGENCY STAFF** |  |  |
| **WEEKDAY** |  |  |
| MORNING | 15% | 36% |
| AFTERNOON | 9% | 5% |
| EVENING/NIGHT | 45% | 44% |
| **WEEKEND** |  |  |
| MORNING | 19% | 43% |
| AFTERNOON | 14% | 14% |
| EVENING/NIGHT | 37% | 39% |

**Table 6.** Registered nurse and nursing support worker costs (£) by Agenda for Change band (substantive and temporary staff)

| AGENDA FOR CHANGE BANDS | SUBSTANTIVE STAFF |  |  | BANK STAFF |  |  | AGENCY STAFF |  |  |
| --- | --- | --- | --- | --- | --- | --- | --- | --- | --- |
|  | Salary a | Employer on cost | Total cost per hour b | Salary | Employer on cost c | Total cost per hour b | Salary d (agency cap maximum) | Employer on cost | Total cost per hour e |
| **1** |  |  |  |  |  |  | 15,516 | 8,534 | 12.29 |
| **2** | 16,536 | 3,548 | 12.63 | 16,536 | 2,359 | 11.88 | 17,978 | 9,888 | 14.24 |
| **3** | 18,333 | 4,054 | 14.08 | 18,333 | 2,736 | 13.25 | 19,655 | 10,810 | 15.56 |
| **4** | 20,279 | 4,602 | 15.65 | 20,279 | 3,144 | 14.73 | 22,458 | 12,352 | 17.79 |
| **5** | 26,038 | 6,225 | 20.48 | 26,038 | 4,353 | 19.3 | 28,462 | 15,654 | 22.55 |
| **6** | 32,342 | 8,002 | 25.62 | 32,342 | 5,676 | 24.14 | 35,225 | 19,374 | 27.9 |
| **7** | 38,801 | 9,822 | 30.87 | 38,801 | 7,032 | 29.1 | 41,373 | 22,755 | 32.77 |
| **8A** | 45,544 | 11,722 | 36.36 | 45,544 | 8,447 | 34.28 | 48,034 | 26,419 | 38.05 |
| **8B** | 54,307 | 14,191 | 43.49 | 54,307 | 10,287 | 41.01 | 57,640 | 31,702 | 45.66 |
| **8C** | 63,703 | 16,839 | 51.14 | 63,703 | 12,259 | 48.23 | 68,484 | 37,666 | 54.25 |
| **8D** | 75,171 | 20,071 | 60.47 | 75,171 | 14,666 | 57.04 | 82,434 | 45,339 | 65.3 |
| **9** | 88,526 | 23,834 | 71.34 | 88,526 | 17,469 | 67.3 | 99,437 | 54,690 | 78.77 |

A mean salary in 2017 as reported in unit costs of health and social care(Curtis and Burns, 2017)
B cost per hour based on annual hours in unit costs of health and social care(Curtis and Burns, 2017): 1,590 (42.4 weeks \* 37.5) for band 2-4 and 1,575 (42 weeks \* 37.5) for band 5 and above
C INCLUDES EMPLOYER NATIONAL INSURANCE CONTRIBUTION AND ASSUMES 50% BANK STAFF INCLUDE EMPLOYER SUPERANNUATION PAYMENT
D SALARY IS UPPER SPINE POINT OF RANGE FOR EACH BAND – METHOD ADOPTED IN AGENCY CAP (NHS IMPROVEMENT, 2018A)
E SALARY PER HOUR BASED ON ANNUAL HOUR CALCULATION FOR AGENCY CAP (NHS IMPROVEMENT, 2018A) – 52.18 WEEKS (365.25/7) \* 37.5 = 1,956.75.

**Table 7.** Coefficients and baseline parameters used in cost-effectiveness models.

| Parameter | Coefficient | Value<sup>a</sup> | LCL | UCL |
| --- | --- | --- | --- | --- |
| Mortality<sup>b</sup> effect [Upper, lower 95% confidence interval] |  |  |  |  |
| Day of Low registered nurse staffing | HR<sup>c</sup> | 1.03 | [1.01, | 1.06] |
| Day of Low nursing support worker staffing | HR | 1.04 | [1.02, | 1.07] |
| Temporary staffing models<sup>d</sup> |  |  |  |  |
| Day of Low registered nurse staffing | HR | 1.03 |  |  |
| Day of Low nursing support worker staffing | HR | 1.05 |  |  |
| Day of High temporary registered nurse staffing | HR | 1.12 |  |  |
| Day of High temporary nursing support worker staffing | HR | 1.05 |  |  |
| Baseline Mortality Rate<sup>e</sup> |  |  |  |  |
| Hospital Trust A | % | 3.21 |  |  |
| Hospital Trust B | % | 3.87 |  |  |
| Hospital Trust D | % | 3.15 |  |  |
| Length of stay effect<sup>b</sup> |  |  |  |  |
| Average registered nurse staffing relative to mean | $\beta$ (raw) | -0.23 | | |
| Average nursing support worker staffing relative to mean | B (raw) | 0.076 |  |  |
| Mean Length of Stay (in 5 days)<sup>f</sup> |  |  |  |  |
| Hospital Trust A | days | 3.65 | (2.89) |  |
| Hospital Trust B | days | 6.33 | (3.74) |  |
| Hospital Trust D | days | 5.05 | (3.15) |  |
| Cost of excess bed day<sup>g</sup> |  |  |  |  |
|  | £ | 337 |  |  |

<sup>a</sup> Values reported here are rounded, however for the modelling reported here, we used precise values as originally calculated.
<sup>b</sup> Derived from Griffiths et al. (2018)
<sup>c</sup> HR Hazard Ratio
<sup>d</sup> As the temporary staffing models also included the effect of low staffing in the original analysis we used the low staffing coefficients from the model that generated the temporary staffing coefficients, hence the slight differences.
<sup>e</sup>NHS Digital (2018b)
<sup>f</sup>NHS Digital (2018a)
<sup>g</sup>NHS Improvement (2018b)

**Table 8.** Changes in Costs, effects and cost-effectiveness for resilient and flexible establishments relative to ‘standard’ establishments for varying levels of temporary staff availability without the use of float staff.

| Temporary staff availability | Staff cost |  | Hospital stay |  | Death |  | NNT (NNH) |  | Staff cost / life |  | Net cost / life |  |
| --- | --- | --- | --- | --- | --- | --- | --- | --- | --- | --- | --- | --- |
|  | Establishment approach |  | Establishment approach |  | Establishment approach |  | Establishment approach |  | Establishment approach |  | Establishment approach |  |
|  | High (Resilient) | Low (Flexible) | High (Resilient) | Low (Flexible) | High (Resilient) | Low (Flexible) | High (Resilient) | Low (Flexible) | High (Resilient) | Low (Flexible) | High (Resilient) | Low (Flexible) |
| None | 7.8% | -19.9% | -1.4% | 2.4% | -4.2% | 13.5% | 762 | (219) | £40,161 | -£29,842 | £20,404 | -£19,976 |
| Limited | 5.7% | -11.0% | -1.2% | 1.7% | -4.5% | 8.7% | 660 | (357) | £26,654 | -£28,901 | £11,983 | -£17,768 |
| Higher | 4.2% | -5.7% | -1.0% | 0.8% | -3.8% | 4.7% | 828 | (690) | £23,359 | -£28,449 | £10,488 | -£18,277 |
| Unlimited | 3.2% | -2.2% | -0.9% | 0.3% | -3.2% | 2.3% | 1106 | (2372) | £21,375 | -£16,236 | £7,670 | -£7,057 |

**Table 9:**
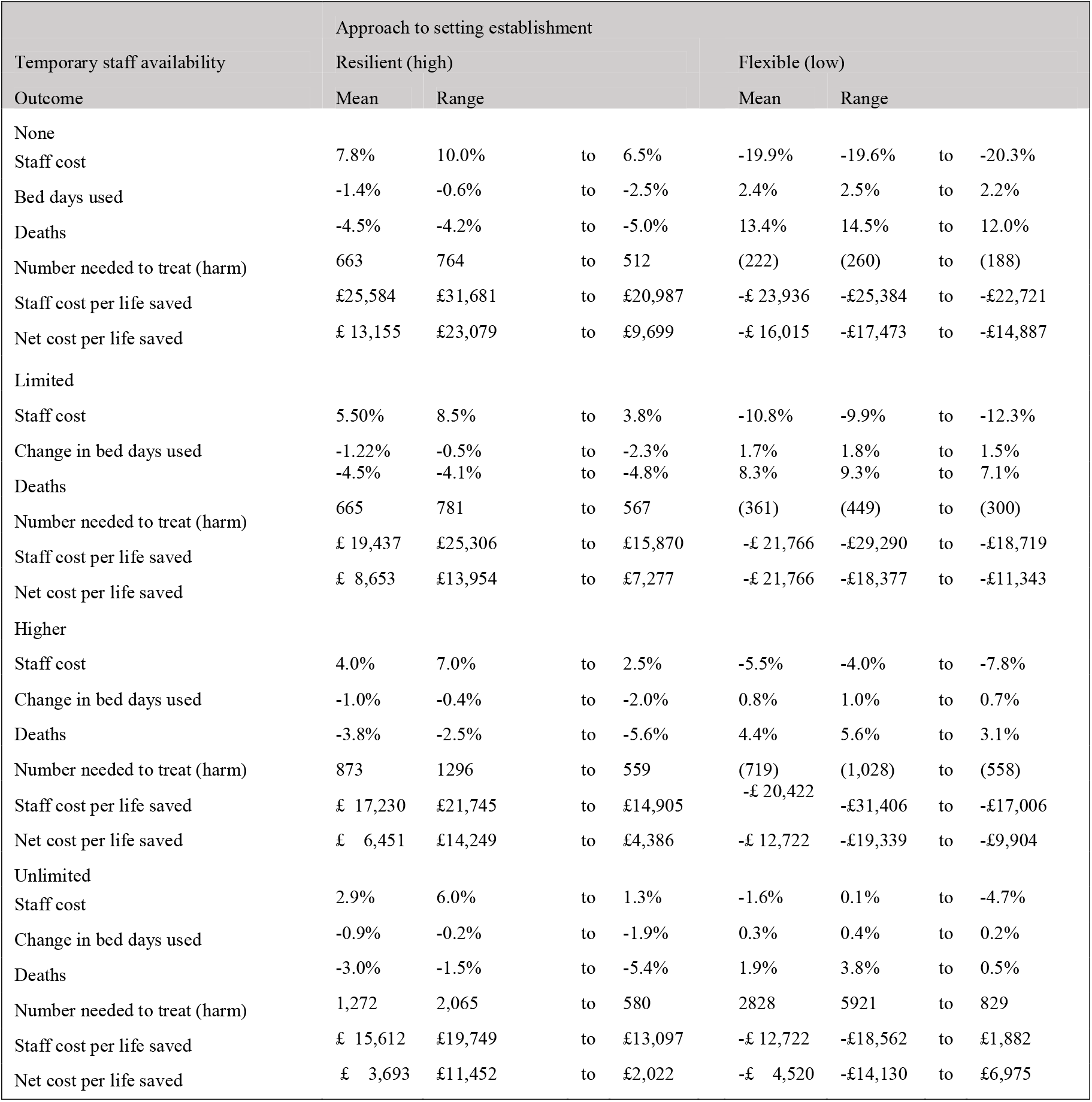
Changes in Costs, effects and cost-effectiveness for resilient and flexible establishments relative to ‘standard’ establishments for varying levels of temporary staff availability (mean and range across three hospital models)

